# Automated Assessment of Choroidal Mass Dimensions Using Static and Dynamic Ultrasonographic Imaging

**DOI:** 10.1101/2025.07.31.25332514

**Authors:** Noah Emmert, Gideon Wall, Amin Nabavi, Amir Rahdar, Matthew Wilson, Benjamin King, Linda Cernichiaro-Espinosa, Siamak Yousefi

## Abstract

**Purpose:** To develop and validate an artificial intelligence (AI)-based model that automatically measures choroidal mass dimensions on B□scan ophthalmic ultrasound still images and cine loops.

**Design:** Retrospective diagnostic accuracy study with internal and external validation.

**Participants:** The dataset included 1,822 still images and 283 cine loops of choroidal masses for model development and testing. An additional 182 still images were used for external validation, and 302 control images with other diagnoses were included to assess specificity

**Methods:** A deep convolutional neural network (CNN) based on the U-Net architecture was developed to automatically measure the apical height and basal diameter of choroidal masses on B-scan ultrasound. All still images were manually annotated by expert graders and reviewed by a senior ocular oncologist. Cine loops were analyzed frame by frame and the frame with the largest detected mass dimensions was selected for evaluation.

**Outcome Measures:** The primary outcome was the model’s measurement accuracy, defined by the mean absolute error (MAE) in millimeters, compared to expert manual annotations, for both apical height and basal diameter. Secondary metrics included the Dice coefficient, coefficient of determination (R^2^), and mean pixel distance between predicted and reference measurements.

**Results:** On the internal test set of still images, the model successfully detected the tumor in 99.7% of cases. The mean absolute error (MAE) was 0.38 ± 0.55 mm for apical height (95.1% of measurements <1 mm of the expert annotation) and was 0.99 ± 1.15 mm for basal diameter (64.4% of measurements <1 mm). Linear agreement between predicted and reference measurements was strong, with R^2^ values of 0.74 for apical height and 0.89 for basal diameter. When applied to the control set of 302 control images, the model demonstrated a moderate false positive rate. On the external validation set, the model maintained comparable accuracy. Among the cine loops, the model detected tumors in 89.4% of cases with comparable accuracy.

**Conclusion:** Deep learning can deliver fast, reproducible, millimeter□level measurements of choroidal mass dimensions with robust performance across different mass types and imaging sources. These findings support the potential clinical utility of AI-assisted measurement tools in ocular oncology workflows.

## INTRODUCTION

Choroidal masses encompass a spectrum of benign and malignant lesions, yet across most of these spectrum tumor dimensions are the strongest single predictors of visual prognosis, therapeutic eligibility, and, in malignant cases, metastatic risk.

Historical data from the Collaborative Ocular Melanoma Study and subsequent series have repeatedly shown that each additional millimeter of basal diameter or height significantly worsens survival and vision outcomes in choroidal malignant melanoma, underscoring the clinical imperative for precise, reproducible measurement (1,2). Although much of the emphasis is on choroidal malignant melanoma (CMM), accurate sizing is also critical for differentiating indeterminate melanocytic lesions, monitoring benign nevi, and planning therapy for metastatic lesions (3,4).

In routine practice, choroidal mass dimensions measured with B-scan ultrasonography, often supplemented by A-scan for acoustic characterization. These examinations demand highly trained ocular sonographers, a workforce that is limited even in tertiary centers, and they remain operator-dependent, contributing to inter-observer variability and access barriers (5).

Artificial intelligence has begun to close similar gaps in other ophthalmic modalities; deep learning systems now compete experts in segmenting choroidal melanomas on fundus photographs, and classifying nevi versus small CMM, yet very few studies have tackled automated choroidal mass dimensions measurement in ocular B-scan ultrasound itself (6–9). By contrast, automated measurement of abdominal lesions on general ultrasound is already well-developed, confirming the feasibility of extracting quantitative metrics (10,11). In this study, we extend these advances to ophthalmology by training a deep learning model that ingests both still B-scan images and cine clips to localize choroidal tumors and output basal diameter and apical height automatically.

## METHODS

### Subjects

We conducted a single center, retrospective cohort study at Hamilton Eye Institute, reviewing patients who attended the ocular oncology clinic with a documented choroidal mass and underwent ophthalmic ultrasonography between 2017 and 2021. A control group was assembled from individuals imaged for other ocular indications (for example, retinal detachment or vitreous haemorrhage) or flat choroidal mass. The Institutional Eligible participants were adults (≥18 years) with a choroidal mass, whether treatment-naïve or previously treated. We excluded still images that lacked paired measurement overlays image, images containing duplicate height or basal-diameter callipers in the same image that indicated highly multilobulated tumours, and any cine loop that did not have a corresponding measured still image. Review Board approved the protocol and waived the requirement for informed consent because of the study’s retrospective design.

Two ultrasound platforms were employed during the study period: the 10-MHz B-scan Eye Cubed system (Ellex Medical Lasers, Adelaide, Australia) from 2017 to 2020 and the 20-MHz B-scan ABSolu platform (Quantel Medical, Cournon-d’Auvergne, France) from 2020 to 2021. Guided by lesion location on colour fundus photography, a single ocular sonographer targeted each tumor from the opposite clock hour in both longitudinal and transverse planes, selecting the frame that best delineated the apex and margins; tumor limits were verified in real time with an ocular oncologist. Still images (.PNG files) acquired on the Eye Cubed constituted the development dataset and were divided patient-wise into training (64 %), validation (16 %), and internal test (20 %) subsets, whereas all images from the ABSolu were reserved as an independent external validation cohort. Cine loops were captured in the same two planes, 9–11 s in length at 30 frames per second, and stored as .WMV files. Cine data were available only from the Eye Cubed system and were reserved solely for test set evaluation. A separate set of still images from the Eye Cubed system, with diagnoses of flat choroidal mass or other than choroidal mass, was used as the control group.

Demographic variables and clinical data were extracted from electronic health records. For each eye we recorded the tumor diagnosis, treatment status (naïve vs. treated), treatment modality, and the interval between treatment and the index ultrasound examination.

### Preprocessing

Still images were exported in pairs of a raw B-scan and its clinician-annotated counterpart. Pairs were verified by pixel-wise similarity ≥ 99.9 % and automatically de-identified. Using simple contour detection and morphological filtering we cropped the region of interest, resized every image to 512 × 512 pixels, and converted them to tensors; horizontal flips, mild affine transforms, and colour jitter were applied for augmentation. Cine loop files were parsed with OpenCV, extracting every fifth frame to limit redundancy and submitting each frame to the identical preprocessing pipeline. Mass measurement lines (basal diameter, apical height) were isolated in HSV color space, after which an ellipse defined by the two-line vectors provided a binary ground truth mask. A per image pixel-to-millimeter factor was derived from the on screen scale bar.

### Deep Learning Model Architecture

Segmentation was performed with a U-Net++ architecture (EfficientNet-B3 encoder) implemented in PyTorch. The network output a single channel logit map that was converted to probabilities with a sigmoid function during training and inference. Optimization used AdamW (initial learning rate 1 × 10□□, weight decay 1 × 10□□) and a compound Dice + binary cross-entropy loss to balance overlap and pixel accuracy. Learning rate was halved after ten stagnant validation epochs; training stopped early after 15 stagnant epochs. A global confidence threshold was calibrated on the validation set (0.1–0.9, step 0.05); 0.20 confidence threshold minimized mean absolute error (MAE) while retaining ≥ 70 % valid masks and was fixed for all test-set inference.

Each preprocessed frame passed through the trained network, producing a probability map that was binarized at the 0.20 threshold. The largest connected component was fitted with a rotated minimum-area rectangle; its long axis yielded basal diameter, the short axis apical height. Frames whose dimensions lay outside a clinically plausible 0.5–50 mm range were discarded. For each video, the frame with the greatest sum of basal diameter and height, empirically the view with optimal probe alignment, was selected for analysis and stored with full metadata.

### Model Evaluation

Segmentation performance was assessed with the Dice coefficient calculated against the ellipse-based pseudo masks. Measurement accuracy was summarized as mean ± SD absolute error (mm), relative error (%), and the coefficient of determination (R^2^) between predicted and ground truth lengths. Concordance rates (< 0.5 mm, < 1 mm, and < 2 mm) were also tabulated, and performance was stratified by lesion size, diagnosis, and treatment status to evaluate model fairness across clinically relevant subgroups. In addition, we performed a pixel-level endpoint analysis; for every image, the Euclidean distance between the predicted and reference coordinates of the two basal diameter endpoints and the two apical height endpoints was computed in native image space, averaged to a mean point distance.

## RESULTS

The main set still image cohort included 1,822 B-scan still images from 871 patients across 1,498 visits. Mean age of the patients was 64.7 ± 14.9 years, of whom 472 (54.2%) were female. The dataset was dominated by CMM and choroidal nevus (89.9% of cases). Overall, 46 % of images were treatment-naïve, while the remainder were obtained a median 2.4 years after therapy. Mean mass height and basal diameter were 2.51 ± 1.90 mm (range: 0.52-15.01 mm) and 8.93 ± 2.79 mm (range: 3.03-21.98 mm), respectively. The control set of stills consisted of 302 images; the test set of Cine loop ultrasound video-matched stills consisted of 283 images; the external validation set includes 302 still images (**Table 1**).

**Table 1.** Patient and image characteristics.

| Variables |  | Main set<br>still images<br>(n=1822,<br>871 pt) | Control set<br>still images<br>(n=302, 27<br>pt) | Main set<br>cine loops<br>(n=283, 216<br>pt) | External<br>validation set<br>still images<br>(n=182, 50 pt) |
| --- | --- | --- | --- | --- | --- |
| **Patients** | **N** | 871 | 27 | 216 | 50 |
| **Visits** | **N** | 1498 | 52 | 250 | 91 |
| **Age (yr)** | **Mean ± SD** | 64.7±14.9 | 60.5±19.1 | 62.1±17.6 | 66.3±12.9 |
| **Sex** | **Male** | 399 (45.8%) | 12 (44.4%) | 103 (47.7%) | 19 (38.0%) |
|  | **Female** | 472 (54.2%) | 15 (55.6%) | 113 (52.3%) | 31 (62.0%) |
| **Diagnosis** | **CMM** | 465 (53.4%) | 6 (22.2%) | 135 (62.5%) | 29 (58.0%) |
|  | **Choroidal**<br>**Nevus** | 318 (36.5%) | 4 (14.8%) | 27 (12.5%) | 21 (42.0%) |
|  | **Choroidal**<br>**Metastasis** | 16 (1.8%) | 0 (0.0%) | 8 (3.7%) | 0 (0.0%) |
|  | **CNV** | 10 (1.1%) | 0 (0.0%) | 7 (3.2%) | 0 (0.0%) |
|  | **Choroidal**<br>**hemangioma** | 8 (0.9%) | 0 (0.0%) | 4 (1.9%) | 0 (0.0%) |
|  | **Others** | 57 (6.5%) | 17 (63.0%) | 35 (16.2%) | 0 (0.0%) |
| **Images** | **N** | 1822 | 302 | 283 | 182 |
| **Treatment**<br>**status** | **Before/No**<br>**treatment** | 839 (46.0%) | 192 (63.6%) | 158 (55.8%) | 62 (34.1%) |
|  | **After**<br>**treatment** | 983 (54.0%) | 110 (36.4%) | 125 (44.2%) | 120 (65.9%) |
| **Time since**<br>**Treatment (yr)** | **Mean ± SD** | 3.8±4.18 | 5.83±1.97 | 3.88±4.76 | 9.08±7.36 |
|  | **Median** | 2.42 | 5.07 | 2.47 | 6.81 |
| **US angle** | **Transverse** | 780 (42.8%) | 101 (33.4%) | 117 (41.3%) | 91 (50.0%) |
|  | **Longitudinal** | 1024<br>(56.2%) | 106 (35.1%) | 166 (58.7%) | 88 (48.4%) |
|  | **Non-targeted** | 18 (1.0%) | 95 (31.5%) | 0 (0.0%) | 3 (1.6%) |
| **Height (mm)** | **Mean ± SD** | 2.51±1.9 | N/A | 4.32±3.12 | 2.0±1.25 |
|  | **Median**<br>**(Range)** | 1.82<br>(0.52-15.01) | N/A | 3.2<br>(0.81-14.79) | 1.5<br>(0.66-6.22) |
|  | **<2.5 mm** | 1296<br>(71.1%) | N/A | 115 (40.6%) | 150 (82.4%) |
|  | **2.5-10 mm** | 505 (27.7%) | N/A | 147 (51.9%) | 32 (17.6%) |
|  | **>10 mm** | 21 (1.2%) | N/A | 21 (7.4%) | 0 (0.0%) |
| **Basal**<br>**diameter**<br>**(mm)** | **Mean ± SD** | 8.93±2.78 | N/A | 11.14±3.97 | 7.54±2.33 |
|  | **Median**<br>**(Range)** | 8.46<br>(3.03-21.98) | N/A | 10.42<br>(3.15-23.27) | 6.97<br>(4.07-14.67) |
|  | **<5 mm** | 68 (3.7%) | N/A | 9 (3.2%) | 153 (84.1%) |
|  | **5-16 mm** | 1754<br>(96.3%) | N/A | 274 (96.8%) | 29 (15.9%) |
|  | **>16 mm** | 38 (2.1%) | N/A | 33 (11.7%) | 0 (0.0%) |

SD: Standard deviation; CMM: Choroidal malignant melanoma; CNV: Choroidal neovascularization

On the development cohort test still images, visual inspection confirmed that the network was spatially aware in 99.7% of images, with both the apical height and basal diameter measurement lines lay on the mass (**Figure 1**). However, in control still images, the model generated false positive measurements in several images. Quantitatively, apical height estimates were highly precise, with a MAE of 0.38 ± 0.57 mm; 76.7 % of heights deviated by < 0.5 mm, 95.1 % by < 1 mm, and 98.4 % by < 2 mm. Basal diameter predictions showed similar fidelity, yielding an MAE of 0.98 ± 1.13 mm; 39.5 % differed by < 0.5 mm, 64.4 % by < 1 mm, and 88.5 % by < 2 mm. Predicted versus true lengths demonstrated strong linear agreement (R^2^ = 0.74 for apical height and 0.89 for basal diameter). Pixel-level endpoint errors, expressed as mean point distance, were 17.51 ± 18.35 pixels for basal diameter and 14.94 ± 17.29 pixels for apical height, while the mean Dice coefficient between predicted masks and pseudo ground truth ellipses was 0.77 ± 0.17 (**Figure 2**).

**Figure 1.**
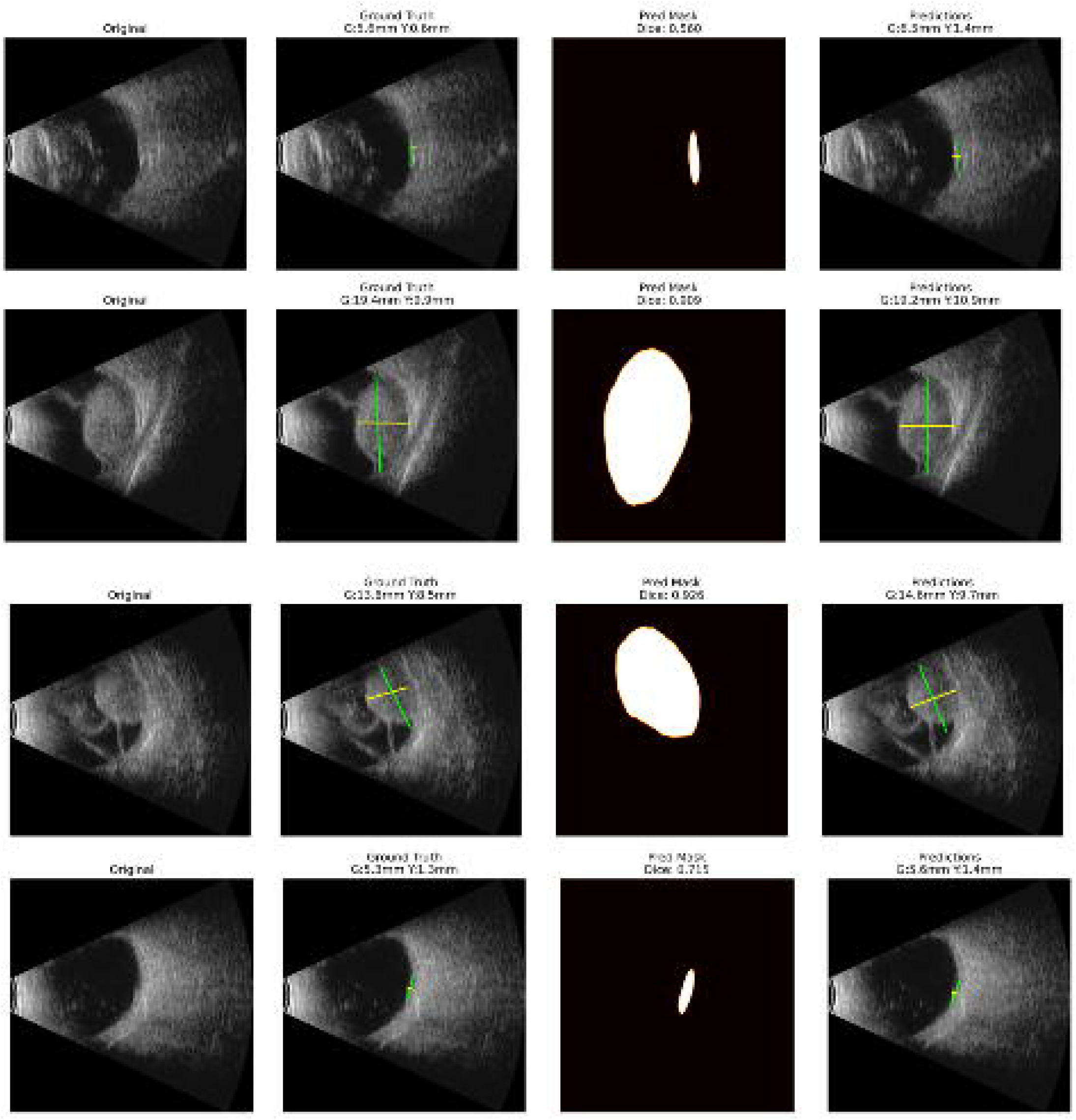
Representative examples of B-scan ultrasound still images from the validation dataset. Left to right columns: (1) Original still image without measurement; (2) Ground truth image with expert-annotated mass dimensions; (3) Predicted segmentation mask with Dice coefficient; (4) Model prediction overlay with corresponding predicted dimensions. Each row shows a separate case with varying mass sizes and prediction accuracy. G: basal diameter (Green line); Y: apical height (Yellow line).

**Figure 2.**
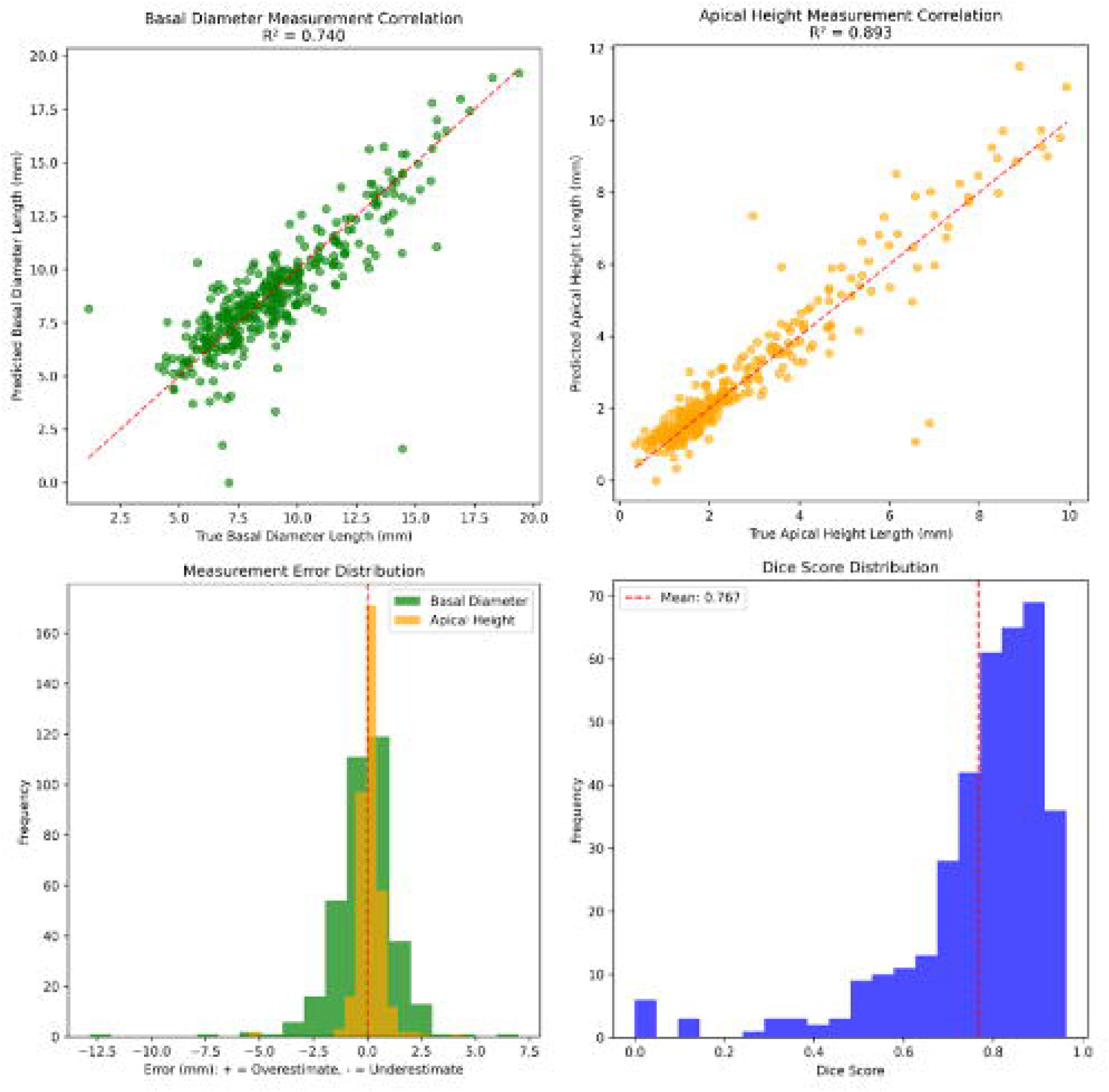
Performance metrics of the deep learning model on the internal test set. Top row: Correlation between predicted and ground truth measurements for basal diameter (left) and apical height (right). Dashed red lines represent the line of best fit. Bottom left: Distribution of measurement error (prediction minus ground truth) for both dimensions, where positive values indicate overestimation. Bottom right: Histogram of Dice similarity scores for mass segmentation.

Model accuracy remained uniformly high across clinically relevant subgroups. Apical height MAE was sub-millimetre for both small (< 2.5 mm, 0.26 mm) and medium (2.5–10 mm, 0.66 mm) tumours, and basal diameter error did not exceed 1 mm regardless of size. T-tests showed no significant differences by diagnosis (choroidal melanoma/nevus vs. others) or treatment status (pre-vs post-therapy), with all P > 0.1.

In the external validation cohort, the model maintained high accuracy. In cine loop analysis, the model could detect the mass in 89.4% of cases with high accuracy.

## DISCUSSION

Our deep learning pipeline delivered measurement accuracy that meets clinically accepted thresholds for B□scan evaluation of choroidal masses. In a test subset of validation still images, MAE was 0.38 mm for apical height and 0.99 mm for basal diameter. These results remained stable across lesion size, diagnostic categories, and treatment status subgroups, and external dataset indicating robust real□world performance.

Automated measurement is already routine in several ultrasound specialties, most notably abdominal applications such as prostate volume estimation, fetal biometry, and aortic screening, where compact CNN models now match expert tracings and can even run on handheld devices (10–12). In ophthalmic ultrasound, however, few published studies have addressed direct, quantitative sizing of intraocular lesions. The gap is clinically important as U.S., as the nationwide supply of diagnostic sonographers and trained ocular oncologists lags demand (13,14). Accurate size assessment is central to treatment selection (e.g., plaque brachytherapy vs enucleation) and to longitudinal surveillance, which ideally uses identical probe planes captured by the same operator. Fully automated sizing could therefore offset staffing shortages, standardize tumor measurements when personnel change, and improve consistency across serial follow-up visits.

Our model was spatially aware in nearly all cases, with apical height error averaged less than 0.5 mm, placing 95 % of measurements within 1 mm of the reference and comfortably below both the interobserver variability reported for human examiners on 10 MHz B-scan (±0.46 mm) and the 0.5 mm change generally considered the minimum threshold for detecting true growth or recurrence (5,15). Together, these findings show that automated sizing can replicate, and in the case of apical height, potentially exceed, expert performance while providing fully reproducible, millimetre-level measurements for clinical decision making and longitudinal follow up.

In this study, we present the first evaluation of an AI model applied to ophthalmic B-scan cine loops. Future work should prospectively test the model on non-targeted “general swipe” cine sweeps of the posterior segment acquired with standardized protocols. Demonstrating robust performance on these untargeted scans would further reduce operator dependence and broaden access to precise, reproducible choroidal mass measurements in settings where specialist imaging expertise is limited.

False positive detections occurred in a subset of control eyes but are acceptable for the intended application. The model is designed for quantitative follow-up of previously identified choroidal lesions, such as post-treatment surveillance of malignant melanoma and longitudinal monitoring of choroidal nevi, which we envision could be performed in general ophthalmology clinics rather than exclusively in ocular oncology centers. In this context, precise size tracking matters more than de-novo lesion detection, and occasional over-segmentation does not compromise clinical decision making. Moreover, the modest tendency toward over-segmentation improves sensitivity for very small masses. No comparable issue was seen in cine loop analysis, because the “best frame” approach inherently excludes non-lesion sweeps.

Several limitations warrant discussion. First, image acquisition still required a trained sonographer to deliberately center and orient the probe toward the tumor. Second, although we performed external validation on an independent device, the same experienced sonographer collected those images, so generalizability remains unproven. Third, our pseudo–ground truth relied on ellipse□based endpoint annotations rather than full volumetric formal tumor segmentation. Ellipses are acceptable for dimension□centric tasks, yet they simplify irregular margins and may embed systematic bias; however, multilobulated choroidal tumors, which are most susceptible to this limitation, are uncommon in routine practice (16), so resulting error is likely minimal; nonetheless, future work should explore pixel-wise annotations to capture complex morphologies more faithfully. Finally, the dataset contained relatively few very large lesions, limiting assessment of the model performance at the extreme end of the size spectrum.

In conclusion, our study demonstrates that deep learning can provide accurate, and reproducible measurements of choroidal mass dimensions from ophthalmic B□scan still images and cine loops. By closing a critical automation gap in ocular ultrasound, the approach has the potential to alleviate expert shortages, enable follow□up assessments in general ophthalmology clinics. Prospective, multicenter evaluations and assessment of “general swipe” B-scan will be the next steps toward clinical translation.

## Data Availability

All data produced in the present study are available upon reasonable request to the authors

